# Mucocutaneous Manifestations of Dengue Fever and Its Correlation with Disease Severity: A Cross-Sectional Study

**DOI:** 10.1101/2025.04.04.25325230

**Authors:** Mohan Bhusal, Sanjay Dhungana, Rabin Baniya, Sulav Wagle, Nikesh Yadav, Shristi Shrestha, Deeptara Pathak Thapa

## Abstract

**Introduction:** Dengue fever (DF), a mosquito-borne viral illness, has seen a rise in cases worldwide including Nepal. While systemic manifestations are well-documented, mucocutaneous lesions are less studied. This study aims to identify the mucocutaneous manifestations of DF and explore their correlation with disease severity, marking the first such effort in Nepal.

**Methods:** This hospital-based descriptive cross-sectional study was conducted between August 2023 to July 2024 at Nepal Medical College Teaching Hospital, a tertiary care center in Kathmandu, Nepal. Ethical approval was obtained from the Institutional Review Committee of Nepal Medical College (IRC Reg. No-11-080/81). Non-probability convenient sampling was used. A total of 151 patients clinically diagnosed with DF and confirmed by NS1 antigen and/or IgM positive tests were included. Data were collected using a structured proforma. Statistical analysis was performed using Statistical Package for Social Sciences (SPSS) version 16.

**Results:** Out of 160,795 patients who visited our hospital from August 2023 to July 2024, 2,059 underwent testing for DF, of which 340 were seropositive, yielding a prevalence of 16.51% (95% CI: 14.97– 18.18) among suspected cases. Among the 151 participants with DF, 93(61.59%) had mucocutaneous involvement, 42 (27.81%) with cutaneous lesions only and 10(6.62%) with mucosal involvement only. Both mucosal and cutaneous manifestations were identified in 41(27.15%). The most common rash was maculopapular 55(36.42%). Pruritus was reported by 65(43.04%) of patients. It was significantly correlated with mucocutaneous involvement (p<0.05). Disease severity classification showed 87 (57.62%) had dengue without warning signs, 52 (34.44%) with warning signs, and 12(7.95%) with severe dengue. Mucocutaneous manifestations was not significantly associated with disease severity in this study.

**Conclusion:** Mucocutaneous manifestations, including maculopapular rashes and pruritus, are common in DF but they do not necessarily correlate with disease severity. Identifying these signs early can improve diagnosis and management.

## Introduction

Dengue is a mosquito-borne viral illness resulting from an infection with one of the four dengue virus serotypes (DENV 1-4), which are transmitted to humans by Aedes aegypti or Aedes albopictus mosquitoes. The infection can range from being asymptomatic or subclinical to fatal shock syndrome.^1^

In 1997, dengue was classified into Dengue Fever (DF), Dengue Hemorrhagic Fever (DHF), and Dengue Shock Syndrome (DSS). In 2009, the WHO revised this to dengue without warning signs, dengue with warning signs, and severe dengue.^2^ The global incidence of dengue has markedly increased over the past two decades making it a serious public health problem.^3^ Nepal has seen a significant rise in dengue cases since 2004, reaching a peak of 54,784 cases and 88 deaths in 2022.^4^ In 2024 till December, 34385 cases of dengue with 13 deaths have been reported.^5^ It is primarily prevalent in the Terai region of Nepal; however, in recent years, its trend has been rising in the hilly areas of the country too.^6^

Mucocutaneous manifestations occur in up to 80% of dengue patients, with skin lesions that may be the first symptom to aid the diagnosis. However, limited research has investigated the clinical significance, complications, and outcomes of skin rashes in dengue infections.^7,8^

This study aimed to identify the prevalence of dengue fever, mucocutaneous manifestations of DF and correlate them with disease severity at a tertiary care center. Up to our knowledge, this is the very first study in Nepal to explore the association between disease severity and mucocutaneous manifestations.

## Methods

This hospital-based cross-sectional study was conducted from August 2023 to July 2024 at Nepal Medical College Teaching Hospital (NMCTH), a tertiary hospital in Kathmandu, Nepal. Total number of patients visiting the hospital and who were tested for dengue antigen during that period were identified using Sukra Hospital Management Information System (SHIMS) version software V1.2.24.5 for prevalence calculation. The study included patients of all ages and genders attending the Outpatient department (OPD), Emergency department, and admitted cases who presented with clinical features suggestive of DF and were dengue virus Non-Structural (NS) 1 antigen and/or IgM positive. Exclusion criteria included patients who lacked signs and symptoms suggestive of DF or tested negative for dengue NS1 antigen and IgM antibody tests, patients with lacking medical records and seronegative dengue cases. A simple convenient, non-probability sampling technique was used.

The required sample size was calculated using the formula:

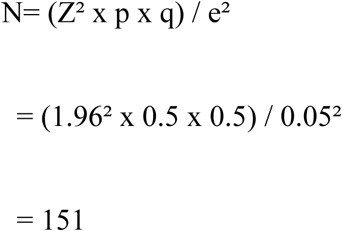

Where:

- N= Minimum required sample size
- Z= 1.96 at a 95% confidence interval (CI)
- p= 50% taken for maximum sample size.
- q= 1 − p = 50%
- e= 8% (margin of error)

The calculated sample size(N) was determined to be 151.

Written informed consent was obtained from all participants aged 16 and above or their guardians for those under 16 years. Anonymity and privacy of patient information were ensured. Data were collected using a preformed proforma via face-to-face interviews and hospital records. A detailed history was recorded, and clinical mucocutaneous and systemic examinations were performed. Data was collected on the demographic profile of the patients, mucocutaneous manifestations and laboratory diagnostic findings, including hematological and biochemical investigations. The proforma included data on age, sex, address, occupation, presenting symptoms and signs, mucocutaneous involvement, cutaneous lesion characteristics (onset, site, morphology, itching, body surface area involvement), mucosal lesions affecting the oral cavity, eyes, or genitalia, and laboratory tests. Complete blood count (CBC) components such as hemoglobin (Hb), hematocrit (Hct), platelet count, white blood cell (WBC) counts were done in all individuals in serologically positive dengue cases. Other laboratory tests like aspartate transaminase (AST), alanine transaminase (ALT), renal function tests (RFT), urine routine and microscopic examination (R/M/E) and radiological investigations were performed to assess disease severity. Thrombocytopenia was defined as a platelet count of less than 150,000/cubic mm of blood, while leukopenia was defined as a leukocyte count of less than 4,000/cubic mm of blood, as per WHO guidelines.^9^

Cases were classified into three groups based on dengue severity according to WHO classification^10^:

1. Dengue without warning signs: Fever plus two of the following symptoms in setting of travel to or residence in endemic area: nausea and vomiting, rash, headache or eye pain or muscle pain or joint pain, leukopenia, and a positive tourniquet test.
2. Dengue with warning signs: Dengue infection plus any of the following symptoms: abdominal pain or tenderness, persistent vomiting, ascites or pleural effusion, mucosal bleeding, lethargy or restlessness, hepatomegaly more than 2 cm, or increased hematocrit with rapid decrease in platelet count.
3. Severe dengue: At least one of the following: severe plasma leakage (shock or fluid accumulation with respiratory distress), severe bleeding, and severe organ involvement AST or ALT more than or equal to 1000 U/L, impaired consciousness, or organ failure).

Data were recorded in a pre-designed proforma and analyzed using SPSS version 16. Descriptive statistics such as frequency, percentages, and mean± standard deviation were calculated and presented in bar diagrams and pie charts. The point estimate was calculated at a 95% Confidence Interval (CI). The chi-square test was applied to assess the correlation between categorical variables, and a p-value <0.05 was considered statistically significant at a 95% CI.

Ethical approval for the study was obtained from the Nepal Medical College Institutional Review Committee (NMC-IRC no-11-080/81). Quality control measures included double verification of data entry by the researcher and thorough analysis of all aspects of the study to ensure accuracy.

The outcome variables were analyzed to evaluate the prevalence of mucocutaneous lesions in dengue patients and their association with disease severity.

## Results

Among the 160,795 patients who visited our hospital between August 2023 and July 2024, 2,059 suspected cases were tested for DF, out of which 340 tested seropositive, indicating a prevalence of 16.51% (95% CI: 14.97–18.18) among suspected cases. Based on the calculated required sample size, we included 151 patients with DF to study mucocutaneous manifestations.

The prevalence of DF was higher among males, 87(57.62%), compared to females 64(42.38%) as shown in figure 1.

**Figure 1.**
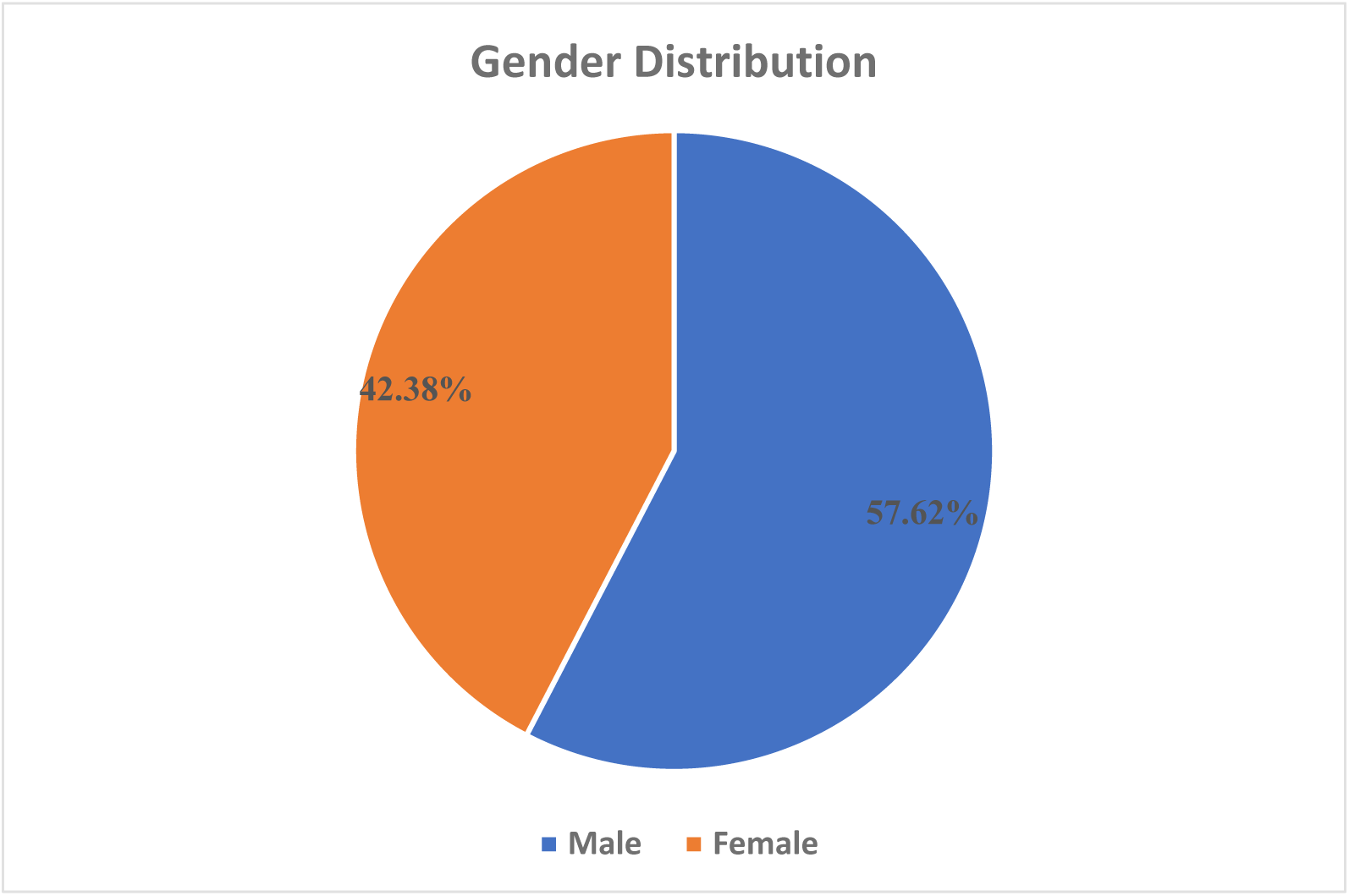
Gender-wise distribution of patients with DF (n= 151) The most commonly affected age group was 21-30 years affecting 39 (25.83%) followed by 31– 40 years age group with 36 (23.84%) (Figure 2).

**Figure 2.**
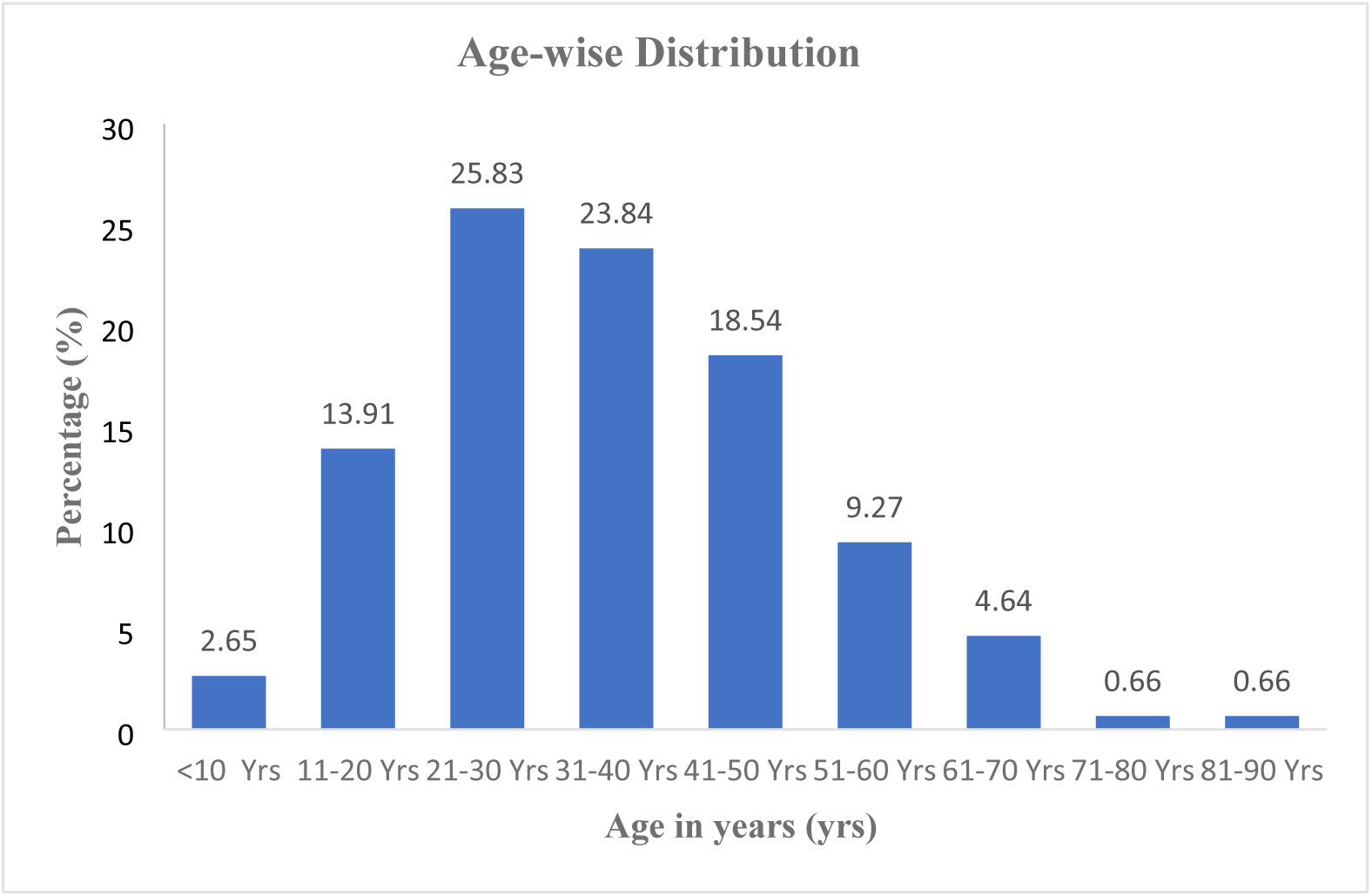
Age group wise distribution of patients with DF (n=151) In our study, mucocutaneous manifestations were observed in 93 (61.59%), as depicted in figure 3. Only cutaneous lesions were present in 42 (27.81%), while mucosal involvement alone was found in 10 (6.62%). Concurrent mucosal and cutaneous manifestations were identified in 41 (27.15%).

**Figure 3.**
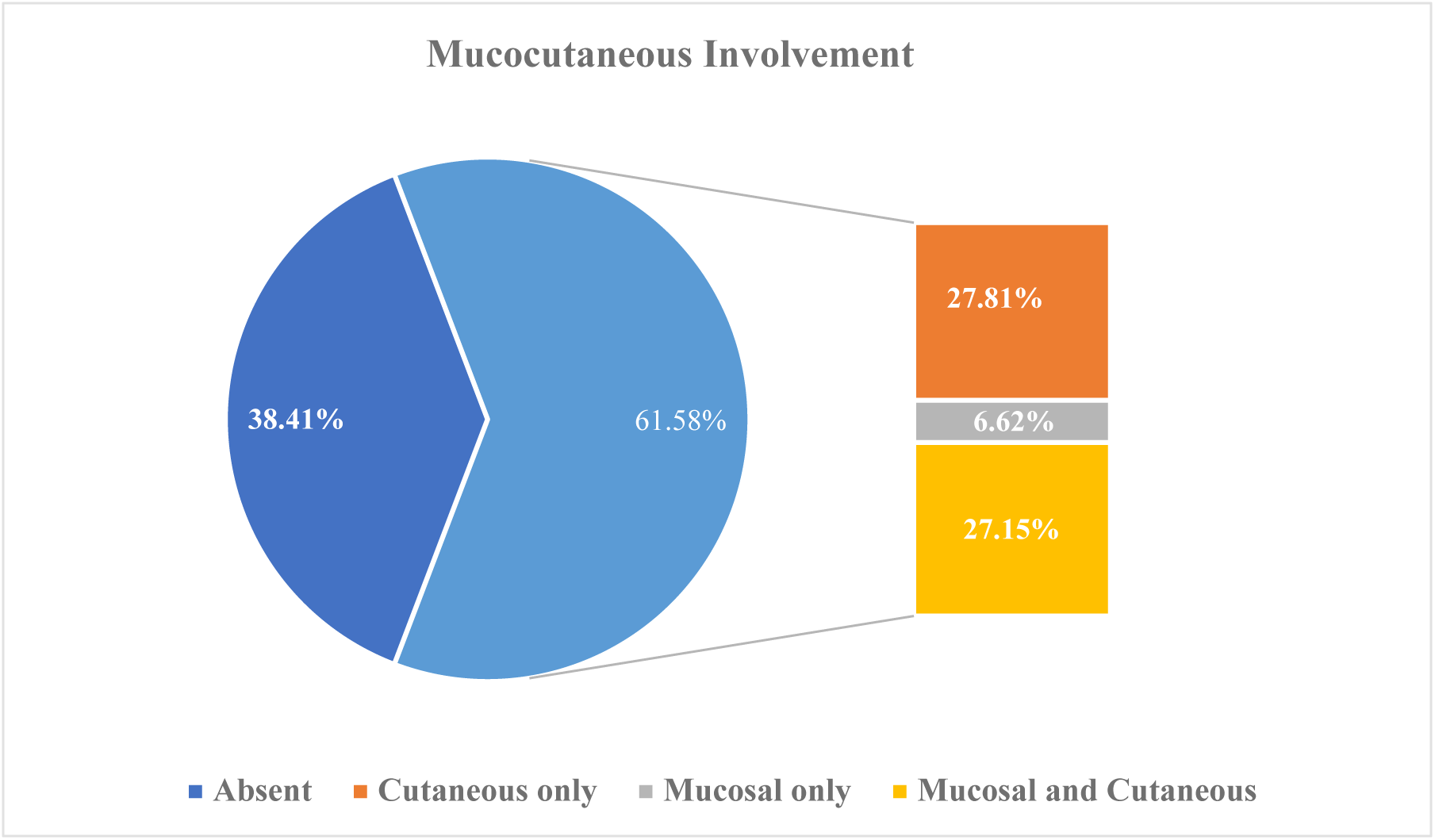
Involvement of mucosal or cutaneous region in patients with DF (n=151) Among mucosal sites, ocular involvement was the most frequent 25 (16.56%), followed by oral cavity lesions 16 (10.60%) and nasal in 10 (6.62%).

Among the observed rash types, maculopapular rash was the predominating type, occurring in 55 (36.42%), followed by erythematous rash in 24(15.89%,) petechiae in 4(2.65%) and no rash in 68(45.03%) (Figure 4).

**Figure 4.**
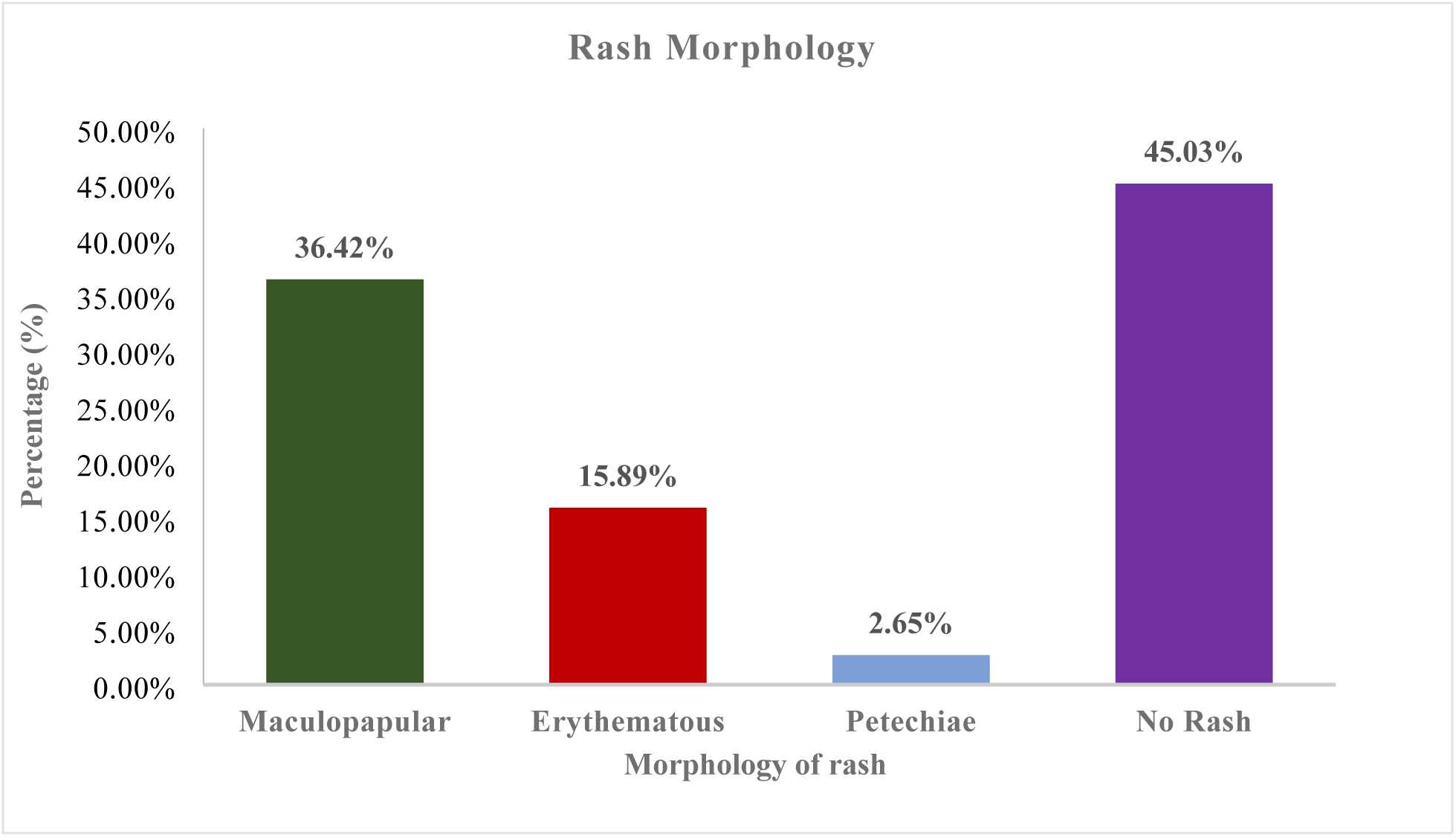
Types of rashes observed in patients with DF (n=151) The onset of rash was most commonly observed between three to seven days, affecting 46(30.46%), with multiple sites being more commonly involved 58 (38.41%) than single sites, and the majority of patients had dengue without warning signs 87(57.6%) followed by dengue with warning signs, 52(34.44%) as shown in Table 1.

**Table 1.** Clinical features of patients with DF.

| Variables | Clinical features of patients with DF | Frequency n (%) |
| --- | --- | --- |
| Onset of Rash, n (83) | < 3 days | 35 (23.18%) |
|  | 3 to 7 days | 46 (30.46%) |
|  | > 7 days | 2 (1.32%) |
| Site, n (83) | Trunk | 9 (5.96%) |
|  | Palms and soles | 5 (3.31%) |
|  | Face | 2 (1.32%) |
|  | Upper limb | 6 (3.97%) |
|  | Lower limb | 3 (1.99%) |
|  | Multiple sites involved | 58 (38.41%) |
| Past History of Dengue, n (151) | Yes | 5 (3.31%) |
|  | No | 146 (96.69%) |
| Platelet counts in cubic mm, n (151) | < 150,000 | 94 (62.25%) |
|  | 150,000-400,000 | 54 (35.76%) |
|  | > 400,000 | 3 (1.99%) |
| WBC counts in cubic mm, n (151) | < 4000 | 55 (36.42%) |
|  | 4000-11,000 | 87 (57.62%) |
|  | > 11,000 | 9 (5.96%) |
| Diagnosis, n (151) | Dengue without warning signs | 87(57.62%) |
|  | Dengue with warning signs | 52(34.44%) |
|  | Severe Dengue | 12(7.95%) |

Itching was the most commonly reported cutaneous symptom, affecting 65 (43.04%) and it showed perfect association with mucocutaneous involvement [odd’s ratio (OR) ∞, (95% CI 16.0-4485)] where all itching cases showed mucocutaneous involvement and also, there were statistically significant higher odds of mucocutaneous involvement among females (OR 2.46, 95% CI 1.23-4.93) than males (p value<0.05) (Table 2).

**Table 2.**
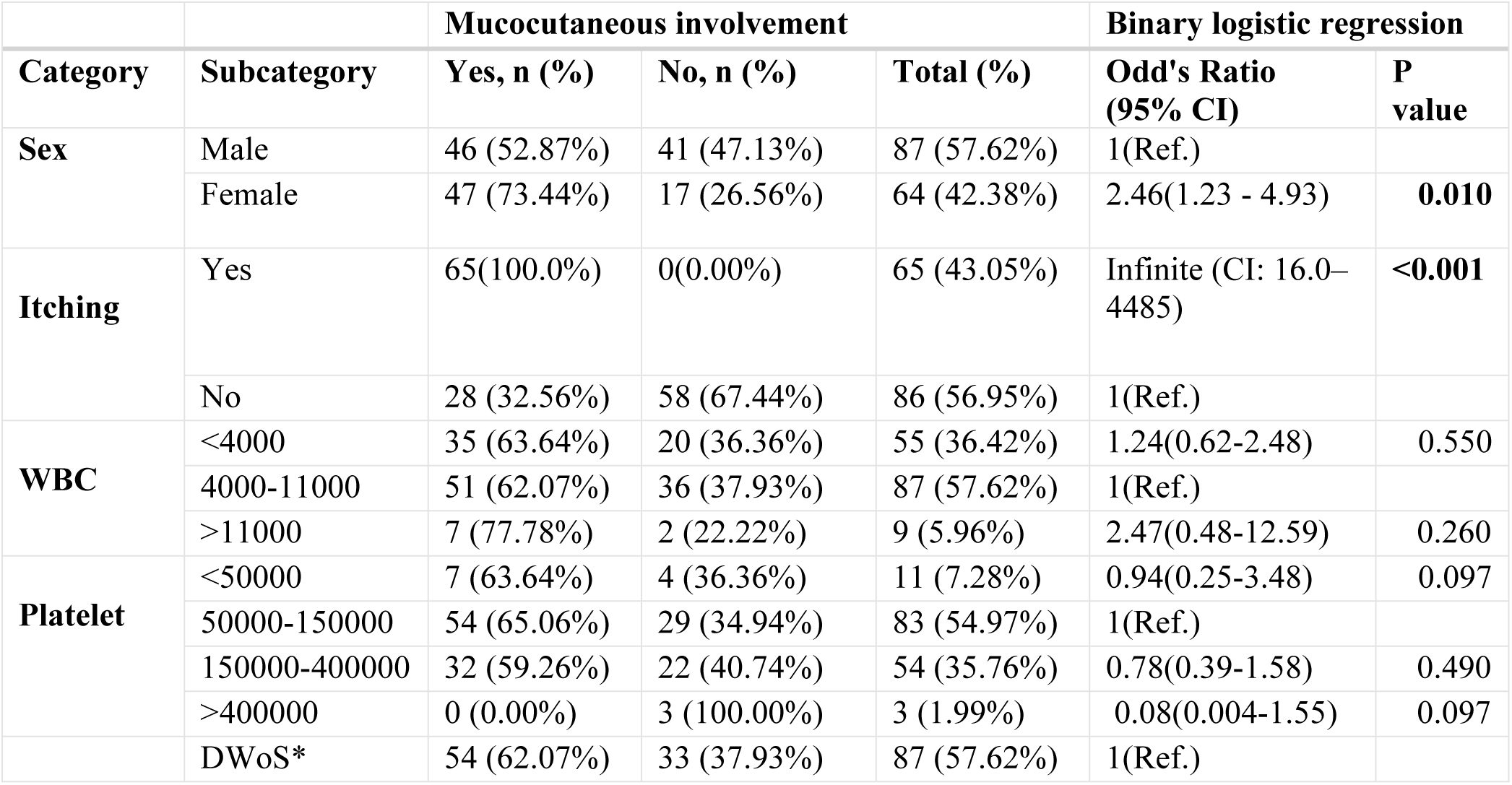
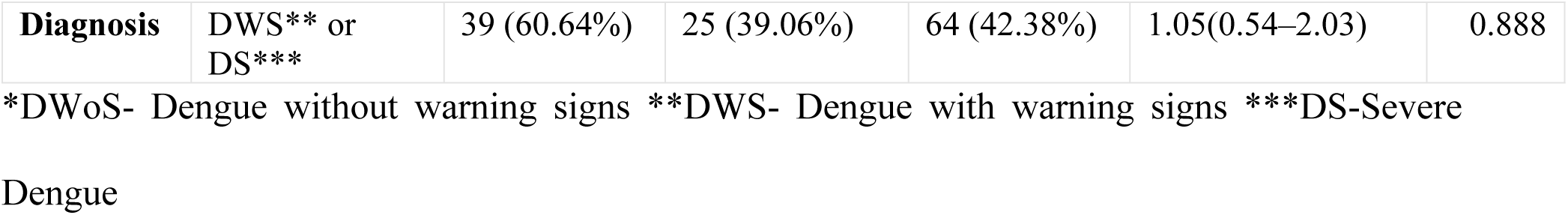
Showing association between mucocutaneous involvement and different parameters in patients with DF (n=151).

## Discussion

Our study showed a prevalence of 16.51% (95% CI: 14.97–18.18) among suspected cases. This contrasts with a study from Nepal reporting a prevalence of 2.72%.^11^ Since this study included only patients visiting the Dermatology department, the true prevalence of DF might have been underestimated. A systematic review of dengue fever in India reported an overall prevalence of laboratory-confirmed dengue infection among clinically suspected patients at 38.3% (95% CI: 34.8%–41.8%).^12^

In our study, the majority of the affected individuals were males 87(57.62%) than female 64(42.38%). Previous research has also documented a higher prevalence of cases among male patients.^11,13–15^ This trend could be attributed to the fact that men are often the primary earners, spending more time outdoors for work-related activities. Additionally, some individuals sleep outside their homes, choosing outdoor settings over indoor ones, which increases their exposure to mosquito bites compared to females. Nevertheless, Females were more prone to developing mucocutaneous manifestations than male which was statistically significant in our study (P value <0.05). In contrast to this, mucocutaneous manifestations in DF were more commonly observed in females than in males, although this difference was not statistically significant in another study.^16^ This could be attributed to higher sensitivity of female skin and some hormonal component making females more prone to allergic skin reactions.^17^

In our study, the most commonly affected age group was 21–30 years, accounting for 39 cases (25.83%), followed by the 31–40 years age group with 36 cases (23.84%). These findings align closely with studies conducted in Pakistan where Ullah observed 64 (28.1%) in 21-30 years age group followed by 31-40 years 53 (23.2%),^18^ Khan reported 145 (48%) were in the age range of 21-40 years^13^ and Saleem also found similar patterns in age group.^19^ This trend may be attributed to the responsibilities typically placed on individuals in this age range, as they are often tasked with earning a livelihood, living away from home, and engaging in outdoor work, which increases their exposure.

In our study, mucosal or cutaneous manifestations were present in 93(61.59%). Of these, only cutaneous lesions were observed in 42(27.81%), while only mucosal involvement was noted in 10 (6.62%). Both mucosal and cutaneous lesions were present in 41(27.15%). In contrast, 58 (38.41%) exhibited no signs of mucosal or cutaneous involvement. Mucocutaneous manifestations of DF have been observed in up to 80% of patients across various studies.^13,20^ Thomas et al. reported skin involvement in 46.8% of patients and mucosal involvement to occur in 15% to 30% of dengue cases in a study involving 124 cases.^8^ In a separate study, Azfar et al. reported cutaneous manifestations in 65% of patients, with mucosal involvement noted in 40.66%.^21^ A study conducted in Karachi found a similar incidence of skin involvement in 68%, as reported by Saleem.^19^ Our findings align with a study from Nepal that reported a rash in 50 (51.1%) DF patients.^22^ However, another study from Nepal observed cutaneous involvement in 180 (89.11%) patients, with mucosal involvement in 58 (28.7%).^11^ The higher findings in later study could be due to inclusion of only Dermatology OPD patients who tend to consult more for the mucocutaneous manifestations.

The most common mucosal manifestations noted in dengue viral infections are conjunctival and scleral injection, small vesicles on the soft palate, erythema and crusting of lips and tongue.^8^ In our study, eyes were mostly affected mucosal site 25 (16.56%) followed by oral cavity 16 (10.60%). In contrast to our finding, oral cavity was most commonly reported mucosal site in other studies.^11,13,21^ this could be attributed to eye involvement causing discomfort and fear whereas oral cavity lesions may go unnoticed. In another similar study from Nepal, mucosal involvement was not seen in patients with DF.^22^ The absence of mucosal lesions in that study is likely attributable to the small sample size, the resolution of lesions before examination, or delayed hospital presentation, during which mucosal manifestations may have already subsided.^22^

Among the types of rashes observed, maculopapular rash was the most common, occurring in 55 (36.42%), followed by erythematous rash in 24 (15.89%) and petechiae in 4 (2.65%). Comparable results were found in study done in University Hospital of Reunion where maculopapular rash was the most common 35 (28.45%).^23^ Study by Aryal et al.^11^ reported maculopapular rash in 70 (34.65%) and morbilliform rash in 69 (34.15%). However, in another study from Nepal, maculopapular rash was reported in 31 (72.1%).^22^ This variation may be due to the smaller sample size of 43 patients in latter study, which may have resulted in a higher proportion of maculopapular rash. The former study included only Dermatology OPD cases. In contrast, our study encompasses both outpatient and inpatient cases, providing a more accurate representation of maculopapular rash prevalence in DF patients. Comparable findings were also noted in a study from Pakistan, where macular rash occurred in 31.7% of cases.^21^

Pruritus was the most commonly reported symptom in our study, affecting 65 (43.04%). This finding aligns with reports from Brazil, where pruritus was observed in 50.5% of patients.^24^ A similar study from Nepal found pruritus to be the most prevalent clinical feature, present in 197 (97.52%).^11^ In another study from Pakistan, generalized pruritus was observed in 135 (69.2%).^21^Also, we found perfectly significant association between pruritus and mucocutaneous involvement in patients with DF (p value<0.001). Like our study, a study from France showed significant association of pruritus with erythematous rash.^23^ DF may present with itching during recovery, potentially serving as a prognostic marker linked to clinical improvement. The mechanisms behind itching could involve viral infection of skin cells and cytokine release.^25^

In DF, the initial rash is a transient flushing erythema of the face, typically appearing shortly before or within the first 24 to 48 hours of symptom onset. This rash is thought to result from capillary dilation. The second rash typically emerges three to six days after the fever begins and is characterized by an asymptomatic maculopapular or morbilliform eruption.^26^ Similarly, in our study, the onset of the rash was most commonly observed between three to seven days, affecting 49(30.46%). The skin rash reflects a reaction of blood vessels to cytokines, leading to inflammatory cell infiltration and dermal edema due to increased vessel permeability, which occurs as part of an intact immune system response.^8^

Dengue with warning signs along with severe dengue was observed in 64 (42.38%), which is similar to a study in Bangladesh reporting 164 (51.4%) of cases with warning signs including severe dengue as well.^27^ However, it contrasts with a study from Nepal where only 95 (27.3%) exhibited warning signs or severe dengue.^6^ This difference may be due to the absence of radiological findings and detailed clinical examinations, as well as the exclusion of findings that could have developed later in the latter study.

The prevalence of mucocutaneous manifestations in dengue and their relationship with severe dengue varies across studies.^23^

We found non significantly higher odds of mucocutaneous involvement in DF with warning signs or severe dengue [OR 1.05 (95% CI: 0.54–2.03)] than DF without warning signs. This can be compared with one of the studies conducted in children, where mucocutaneous manifestations were more commonly observed in severe dengue compared to DF with warning signs and DF without warning signs, and this was statistically significant (p < 0.05).^16^

A retrospective study involving 847 RT-PCR-confirmed dengue patients found no significant association between mucocutaneous signs and the development of severe dengue (p = 0.447).^24^ Similarly, a prospective study of 163 dengue cases reported no link between mucocutaneous manifestations and severe dengue fever (p = 0.54).^28^ Furthermore, a study conducted in Taiwan observed no association between skin rash and dengue hemorrhagic fever (p = 0.406).^7^

This study represents the first effort in Nepal to explore the relationship between DF and mucocutaneous manifestations. Significant associations of mucocutaneous involvement were observed with gender, presence of pruritus but the findings did not reveal any significant association between this condition and the progression to severe dengue. Additional multicentric studies with larger and more consistent sample sizes are recommended to better understand mucocutaneous manifestations that may potentially be linked to severity of dengue.

This study has few limitations. It is a single-center study, and the serological types of dengue were not investigated. The differentiation between primary and secondary dengue infections was also not performed. Additionally, the limited sample size reduces the statistical power and representativeness of the results. Convenience sampling may not adequately represent the target population, potentially introducing bias into the results. Furthermore, the study’s location, being confined to a single hospital, restricts the generalizability of the findings to a broader population. Future studies with larger, multicenter cohorts are needed to address these limitations.

## Conclusion

Mucocutaneous manifestations, including maculopapular rashes and pruritus, are common in DF, affecting a significant portion of patients. While these manifestations serve as important diagnostic markers, they do not correlate with disease severity. Early identification of skin and mucosal involvement can enhance patient management and improve clinical outcomes as DF carries the risk of progressing to life-threatening complications.

## Data Availability

All relevant data are within the manuscript and its Supporting Information files

## Notes

### Competing Interest Statement

The authors have declared no competing interest.

### Funding Statement

The author(s) received no specific funding for this work.

### Author Declarations

Ethical approval was obtained from the Institutional Review Committee of Nepal Medical College (IRC Reg. No- 11-080/81).

